# Molecular Detection and High-Frequency Horizontal Gene Transfer of ESBL Genotype from *Proteus* Species to *Escherichia coli*: Implications for the Spread of WHO Priority Pathogens in North-Eastern Nigeria

**DOI:** 10.64898/2026.04.29.26352026

**Authors:** Isyaka M. Tom, H. B. Ali, Aishatu A. Ibrahim, Usman A. Dutsinma, Muhammad M. Ibrahim, Askira M. Umoru, Habiba J. Balla

## Abstract

**Background:** The rise of antimicrobial resistance (AMR) in the Lake Chad Basin poses a significant threat to global health. While Escherichia coli and Klebsiella pneumoniae are primary concerns for the WHO, Proteus species have emerged as important clinical pathogens and potential reservoirs for genetic resistance. This study aimed to analyze the molecular diversity and horizontal gene transfer (HGT) potential of ESBL-producing Proteus species in the region.

**Methods:** A regional surveillance was conducted with 1,500 clinical samples from Borno, Adamawa, Bauchi, Gombe, Taraba, and Yobe states. Proteus isolates were identified biochemically, and antibiotic susceptibility was assessed using the Kirby-Bauer method. Resistance genes (blaTEM, blaSHV, blaCTX-M) were identified via PCR, and HGT was evaluated through conjugation assays.

**Results:** A total of 144 Proteus isolates were identified, with a prevalence of 9.6%. P. mirabilis was the dominant species (90.97%). Phenotypic screening indicated that 69.44% produced extended-spectrum beta-lactamases (ESBL), with high resistance rates observed for Cefotaxime (80.56%) and Ampicillin (84.72%). Alarmingly, resistance to Ertapenem reached 54.86%. Molecular analysis showed blaTEM as the predominant gene (81.69%), and the conjugation assay revealed a high HGT rate of 76.92%, confirming blaTEM acquisition by E. coli.

**Conclusion:** These results indicate that Proteus species in North-Eastern Nigeria are significant reservoirs for genetic resistance, facilitating the spread of ESBL markers. The high frequency of HGT raises concerns about the effectiveness of beta-lactam therapies in sub-Saharan Africa, underscoring the need to include Proteus in the GLASS framework and promote regional antimicrobial stewardship efforts.

**Current Understanding:** Antimicrobial resistance (AMR) in Enterobacteriaceae, particularly with Escherichia coli and Klebsiella pneumoniae, is a significant global issue highlighted by the World Health Organization’s Global Antimicrobial Resistance and Use Surveillance System (WHO GLASS). While Proteus species are recognized as opportunistic pathogens, their role as genetic reservoirs in sub-Saharan Africa, especially in the Lake Chad Basin, remains inadequately defined in surveillance data.

**Study Contribution:** This study identifies Proteus species as a critical “Genetic Hub” for the transmission of extended-spectrum beta-lactamases (ESBL) in North-Eastern Nigeria, revealing a high horizontal gene transfer (HGT) rate of 76.92% for the blaTEM genotype to E. coli. It also shows a concerning 54.86% resistance rate to Ertapenem, underscoring the urgent need to include Proteus in regional stewardship and global surveillance efforts.

## INTRODUCTION

The AMR crisis threatens decades of medical advancements, prompting the WHO to implement the Global Antimicrobial Resistance and Use Surveillance System. Despite this, many resource-limited areas remain underreported, particularly regarding the prevalence of ESBLs among Enterobacteriaceae. Proteus species, notably Proteus mirabilis, are increasingly recognized as multidrug-resistant pathogens linked to complex infections, complicating treatment options due to resistance mediated by blaTEM, blaSHV, and blaCTX-M genes.

Horizontal Gene Transfer (HGT) enables the rapid spread of resistance genes, with Proteus acting as a major genetic reservoir for transferring ESBL genotypes to pathogens like E. coli. In the Lake Chad Basin, unique socio-ecological factors exacerbate AMR dynamics, yet there’s a lack of comprehensive regional data on Proteus-mediated resistance conforming to GLASS standards. This study aims to fill this gap by providing extensive surveillance of ESBL-producing Proteus species across six states, clarifying their role in the spread of multidrug resistance.

## 2. MATERIALS AND METHODS

### 2.1 Bacteriological Analysis and Identification

#### 2.1.1 Primary Culture and Isolation

Clinical samples were processed and cultured using standard microbiological methods (Cheesbrough, 2006; Baker et al., 2007). Samples were plated on 5% Blood agar, MacConkey agar, and Cystine Lactose-Electrolyte-Deficient (CLED) agar, followed by incubation at 37°C for 18 hours. Presumptive Proteus colonies were subcultured for pure culture isolation. MacConkey agar served as an inhibitory medium when swarming motility interfered.

#### 2.1.2 Morphological and Biochemical Characterization

Isolates were screened for swarming motility and determined to be Gram-negative bacilli via Gram staining. Definitive identification involved biochemical tests for phenylalanine deamination, urease activity, H2S production, and indole production, alongside Methyl Red and Voges-Proskauer tests.

#### 2.1.3 Standardization and Quality Control

Media were prepared as per manufacturer’s instructions, validated with quality control strains recommended by WHO. Proteus mirabilis ATCC 12453 and E. coli ATCC 25922 served as positive and negative controls.

### 2.2 Antibiotic Susceptibility Testing (AST)

#### 2.2.1 Preparation and Standardization of Inoculum

Pure Proteus isolates were subcultured on agar plates, then suspended in sterile saline and standardized to a 0.5 McFarland turbidity (1.5 × 10^8 CFU/mL) (Ejikeugwu, 2023).

#### 2.2.2 Determination of Antimicrobial Profiles

Antibiotic susceptibility was assessed using the Kirby-Bauer disc diffusion method, with a panel of 22 antibiotics selected based on local availability and WHO AWaRe classification. Incubation at 37°C for 24 hours allowed for measuring zones of inhibition according to CLSI (2023) standards.

### 2.3 ESBL Screening and Phenotypic Confirmation

#### 2.3.1 Presumptive Screening

ESBL screening followed CLSI guidelines, using Ceftriaxone, Ceftazidime, and Cefotaxime. Isolates with specific zone diameters were classified as presumptive ESBL producers.

#### 2.3.2 Phenotypic Confirmatory Test

The Combined Disc Diffusion Method confirmed ESBL production. A lawn culture of MDR Proteus was prepared on MHA, using Ceftazidime discs alone and with clavulanic acid to evaluate zone diameter differences.

### 2.4 Liquid Mating Conjugation Assay

#### 2.4.1 Selection of Strains

To assess Proteus species as a genetic reservoir for antimicrobial resistance, we examined horizontal gene transfer (HGT) using a liquid mating conjugation assay. Only molecularly confirmed ESBL-producing isolates served as donor strains, while a Nalidixic acid-resistant E. coli strain from the University of Maiduguri Teaching Hospital acted as the recipient. Notably, 118 Nalidixic acid-resistant Proteus isolates were excluded from this phase.

#### 2.4.2 Culture and Mating Conditions

Donor isolates were cultured overnight in LB broth with 60 µg/mL Ampicillin, and the recipient strain was grown in antibiotic-free LB broth. For conjugation, 1 mL of each culture was mixed in fresh LB broth and pre-incubated for 2 hours, followed by a 1:1 mixture for 6 hours at 37°C. This setup mimicked the fluid dynamics of the human gastrointestinal tract.

#### 2.4.3 Selection and Validation of Transconjugants

Transconjugants were selected on MacConkey agar (Himedia, UK) with 32 µg/mL Nalidixic acid and 100 µg/mL Ampicillin, based on established protocols. Successful HGT was confirmed by growth on dual-antibiotic media and subsequent phenotypic ESBL confirmation using the combined disc diffusion method.

### 2.5 Molecular Characterization of ESBL Determinants

#### 2.5.1 Plasmid DNA Extraction

Plasmid DNA was extracted from Proteus isolates using the Plasmid MiniPrep Kit (Norgen Biotek Corp, Canada). Colonies from an overnight culture on nutrient agar were inoculated into LB broth and incubated at 37°C. After centrifugation, cells were lysed, and the lysate was clarified. The plasmid DNA was washed, eluted with Buffer K, quantified using NanoDrop, and stored at -20°C.

#### 2.5.2 Primer Design and Validation

Primers for blaTEM, blaSHV, and blaCTX-M genes were adopted and synthesized by Inqaba Biotech, West Africa as shown in supplementary table 1. Their specificity was confirmed using NCBI Primer-BLAST, and they were reconstituted in nuclease-free water to a 100 µM stock and diluted to 10 µM for PCR.

#### 2.5.3 Amplification by PCR

PCR amplification was carried out in 50 μL reactions with 3 μL of template DNA, 1 μL of each primer, and 25 μL of Taq PCR Master Mix. The thermal cycling conditions included initial denaturation, 35 cycles of denaturation, annealing (at specific temperatures), and extension, followed by a final extension step. Target fragment sizes were 868 bp (blaTEM), 865 bp (blaSHV), and 499 bp (blaCTX-M).

#### 2.5.4 Gel Electrophoresis

PCR products were resolved using 1.5% agarose gel electrophoresis in TBE buffer, with ethidium bromide for visualization. Samples were compared to a DNA ladder and visualized using UV trans-illumination.

### 2.6 Ethical and Demographic Reporting

Results were reported by sex and age, following SAGER guidelines. Consent was obtained from participants in accordance with ethics committee protocols.

### 2.7 Statistical Analysis

#### 2.7.1 Data Management

Data from microbiological and molecular assays were entered into Excel and analyzed using SPSS version 26.0.

#### 2.7.2 Inferential Statistics

The Pearson’s Chi-square test was used for categorical variable associations. Evaluations were conducted at a 99% confidence level, with a significance threshold of p < 0.01.

## 3.0 RESULTS

### 3.1. Prevalence and Species Distribution

The initial phase of this study focused on the recovery of Proteus species from various clinical sources across six states in Northeastern Nigeria. As presented in Table 1, a total of 144 isolates were recovered from 1,500 samples, resulting in an overall prevalence of 9.60%. The SSH Maiduguri facility recorded the highest recovery rate at 11.6%, followed closely by FMC Yola at 10.8%. Taxonomically, Proteus mirabilis was identified as the predominant species, accounting for 90.97% (n=131) of the total isolates. Additionally, the rare species P. penneri was identified in Bauchi, contributing to 0.69% of the overall collection.

**Table 1:**
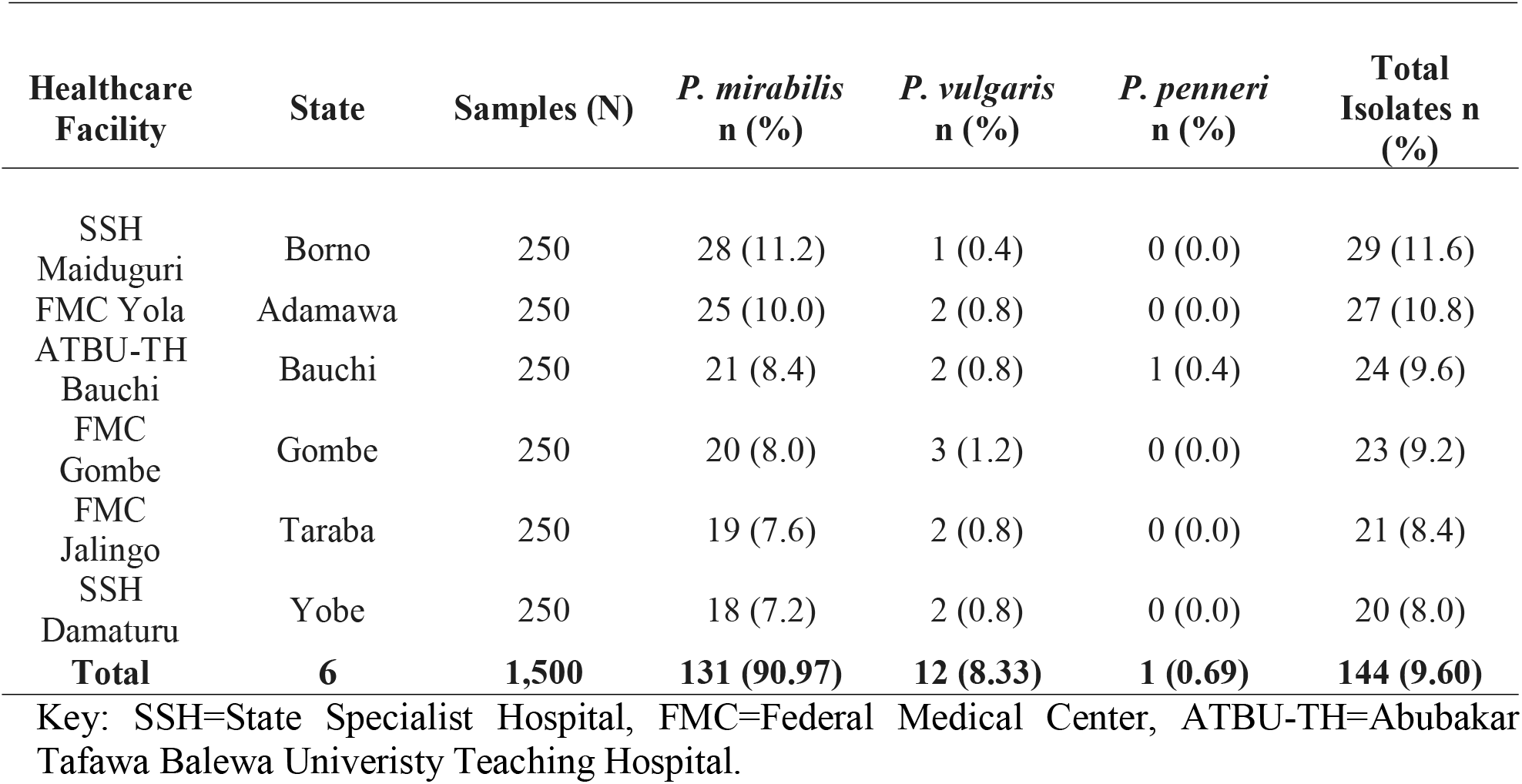
Prevalence and Species Distribution of Clinical Proteus Isolates.

### 3.2. Screening for Beta-Lactamase Phenotypes

Prior to undertaking molecular characterization, all 144 isolates underwent evaluation for the production of beta-lactamase enzymes in order to establish a resistance baseline. The screening results presented in Table 2 demonstrate that 100 out of the 144 isolates (69.44%) were confirmed as beta-lactamase producers. Notably, P. mirabilis accounted for the highest number of producers, with a total of 91. Furthermore, the statistical analysis yielded a P-value of 0.901 (X2 = 0.207), indicating that the production of beta-lactamase is consistently distributed among the genus within the study area, irrespective of the specific species involved.

**Table 2:** Rate of Occurrence of *Proteus* spp producing β-Lactamase in the Study Area.

| *Proteus* spp | Total Isolated (%) | β-lactamase Producing isolates (%) |
| --- | --- | --- |
| *P. mirabilis* | 131 (90.97) | 91 (63.19) |
| *P. vulgaris* | 12 (8.33) | 8 (5.56) |
| *P. penneri* | 1 (0.69) | 1 (0.69) |
| **Total** | **144 (100)** | **100 (69.44)** |

### 3.3. Antimicrobial Susceptibility Profiles

The isolates were subjected to sensitivity testing across various antibiotic classes to assess therapeutic challenges. The resistance profiles presented in Table 3 reveal alarmingly high levels of resistance to beta-lactam antibiotics, with Ampicillin (84.72%) and Cefotaxime (80.56%) exhibiting the most significant susceptibility issues. In contrast, fluoroquinolones demonstrated superior efficacy, particularly Levofloxacin, which maintained a high sensitivity rate of 87.50%. While a resistance rate of 54.86% to Ertapenem was noted among isolates, the P-value of 0.011 suggests that this finding represents a borderline significant trend when compared to the more firmly established resistance observed in other antibiotic classes, thus identifying it as an emerging resistance concern within the sub-region. The difference in resistance rates between beta-lactams and fluoroquinolones was found to be highly significant (P < 0.01), highlighting the substantial erosion of the effectiveness of beta-lactam antibiotics.

**Table 3:**
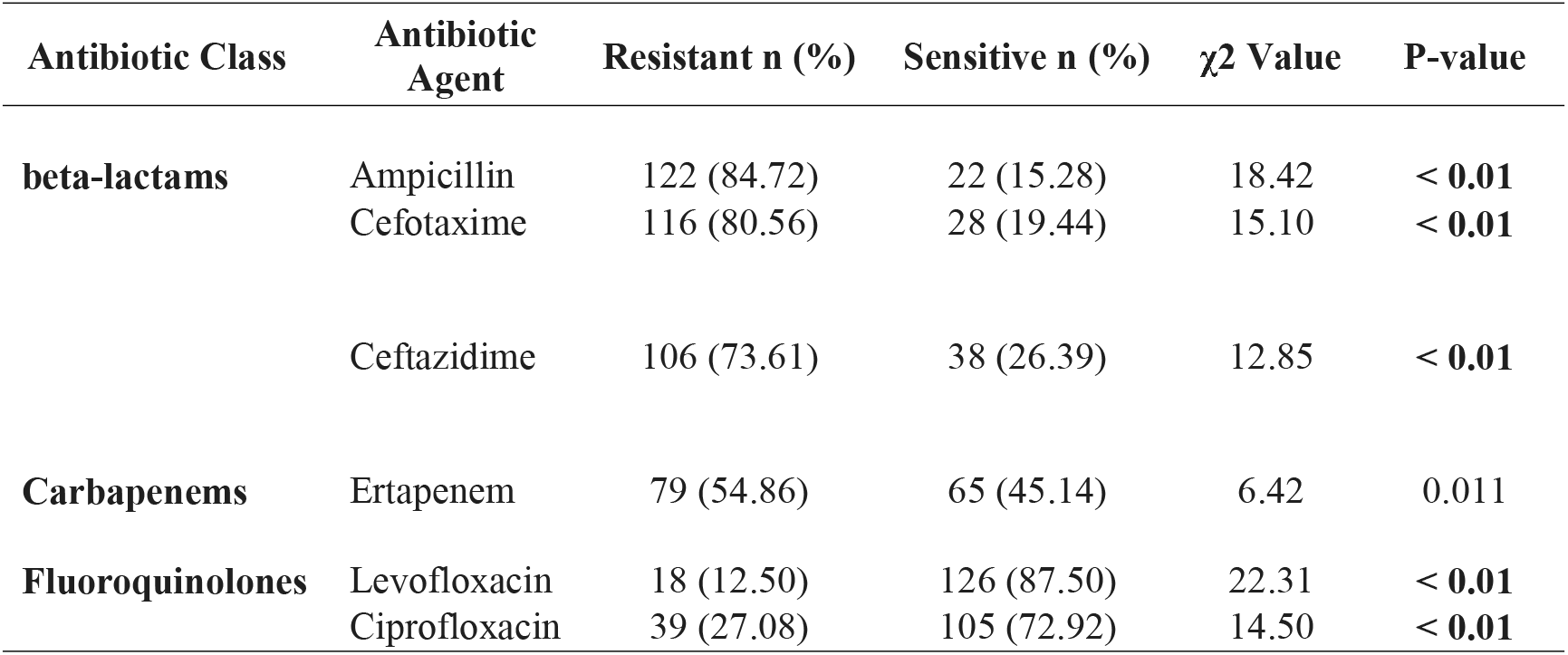
Antimicrobial Resistance Patterns and Statistical Significance.

| Antibiotic Class | Antibiotic Agent | Resistant n (%) | Sensitive n (%) | $\chi^2$ Value | P-value |
| --- | --- | --- | --- | --- | --- |
| beta-lactams | Ampicillin | 122 (84.72) | 22 (15.28) | 18.42 | < **0.01** |
|  | Cefotaxime | 116 (80.56) | 28 (19.44) | 15.10 | < **0.01** |
|  | Ceftazidime | 106 (73.61) | 38 (26.39) | 12.85 | < **0.01** |
| Carbapenems | Ertapenem | 79 (54.86) | 65 (45.14) | 6.42 | 0.011 |
| Fluoroquinolones | Levofloxacin | 18 (12.50) | 126 (87.50) | 22.31 | < **0.01** |
|  | Ciprofloxacin | 39 (27.08) | 105 (72.92) | 14.50 | < **0.01** |

### 3.4. Assessment of Horizontal Gene Transfer Potential

To evaluate the risk associated with the dissemination of resistance, a conjugation assay was conducted to examine the mobility of the R-plasmids. As summarized in Table 4, the outcomes of the conjugation experiments revealed a high success rate of 76.92% (20 out of 26 donors). This significant frequency of horizontal gene transfer indicates that the resistance determinants are present on highly mobile plasmids, thereby posing a substantial risk for the rapid propagation of resistance to other enteric pathogens within the clinical environment Figure 1 (A and B).

**Table 4:** Conjugal Transfer Frequency of ESBL Resistance.

| Parameter | Result |
| --- | --- |
| Total Donors Tested (n) | 26 |
| Successful Transconjugants (n) | 20 |
| Conjugation Success Rate (%) | 76.92% |
| Statistical Power | Significant mobility observed ($P < 0.01$) |

All categorical data were analyzed utilizing the Chi-square test. At a 99% confidence level, the resistance to beta-lactam antibiotics and the distribution of the *bla*TEM gene within the Northeastern sub-region were found to be highly significant ( $P < 0.01$ ), underscoring the clinical urgency of these findings.

**Figure 1A:**
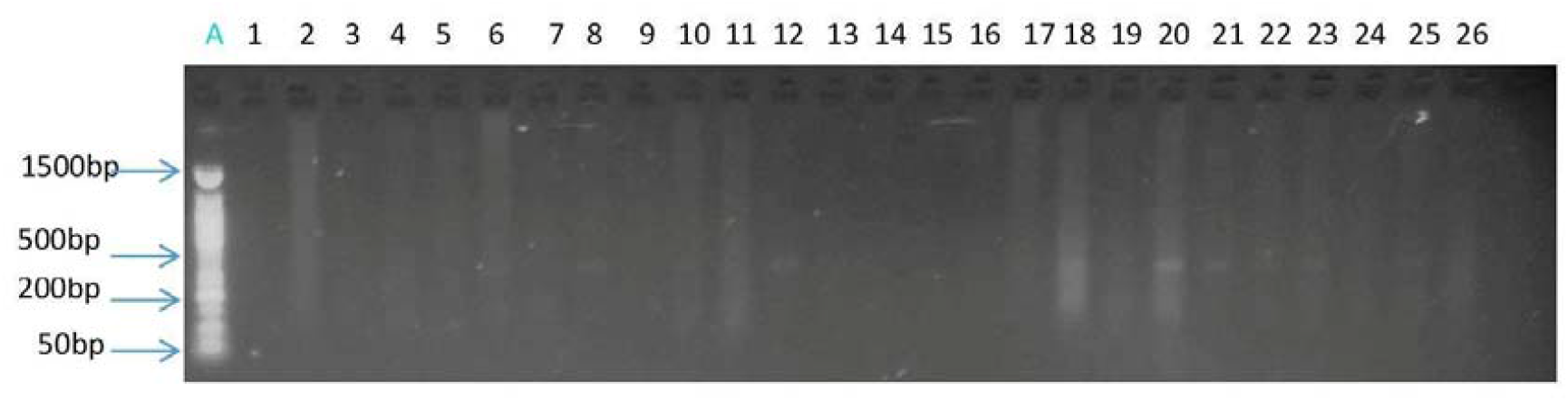
Pre-Conjugation Verification of Recipient Strains. The agarose gel electrophoresis (1.5%) demonstrates the results of PCR amplification for the recipient E. coli J53, which is resistant to Nalidixic acid, prior to the conjugation assay. Lane A contains a 100 bp DNA ladder; Lanes 1–26 exhibit recipient strains that show a complete absence of bla genes. This provides confirmation that the recipient serves as an appropriate negative control for horizontal gene transfer experiments.

**Figure 1B:**
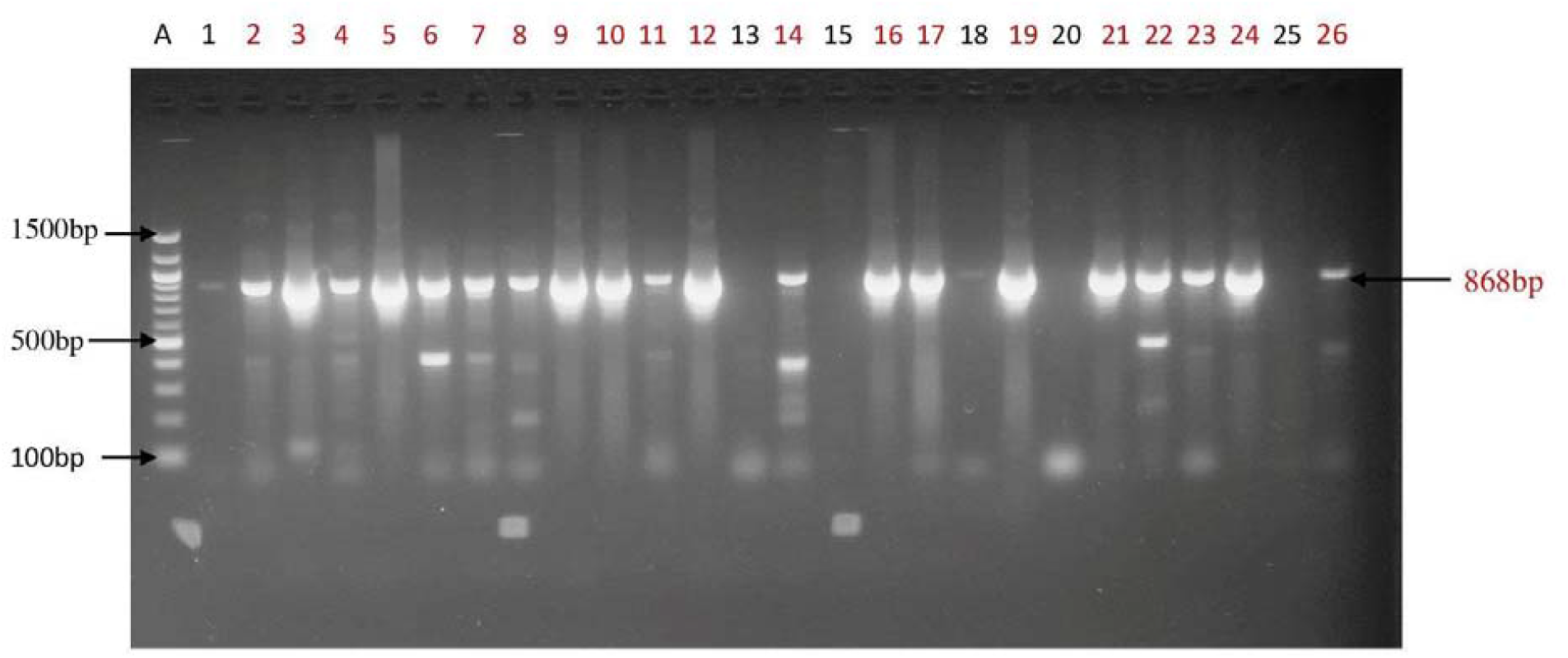
Molecular Confirmation of Transconjugants (blaTEM at 868 bp) The agarose gel electrophoresis (1.5%) displays PCR product results following the conjugation assay. Lane A includes a 100 bp DNA ladder; Lanes 2, 3, 4, 5, 6, 8, 10, 11, 13, 15, 16, 17, 18, 19, 20, 21, 22, 23, 24, and 26 present distinct amplicons at the 868 bp mark, confirming the successful horizontal transfer of the blaTEM gene from the Proteus donors. Conversely, Lanes 1, 7, 9, 12, 14, and 25 depict unsuccessful mating pairs. This visual evidence substantiates the 76.92% phenotypic transfer frequency.

### 3.5. Molecular Detection of ESBL Genotypes

Molecular characterization has identified specific resistance markers in 71% of the phenotypic producers. The distribution of these genes across the six states is detailed in Table 5 and is visually corroborated in Figures 2, 3, and 4.

**Table 5.0:** Distribution of TEM, SHV and CTX-M Genes of ESBL Producing *Proteus* spp. Isolated from the Selected Hospitals.

| Name of Hospital | bla of ESBL Producing Proteus Isolates |  |  |  |  |  |  | Total (%) |
| --- | --- | --- | --- | --- | --- | --- | --- | --- |
|  | Single Gene |  |  | Multiple Gene |  |  |  |  |
|  | TEM | SHV | CTX-M | TEM/SHV | TEM/CTX-M | SHV/CTX-M | TEM/SHV/CTX-M |  |
| SSH Maiduguri | 15<br>(21%) | 0<br>(0.0%) | 0<br>(0.0%) | 1<br>(1.41%) | 0<br>(0.0%) | 0<br>(0.0%) | 0<br>(0.0%) | 16<br>(22.54%) |
| FMC Yola | 8<br>(11.27%) | 0<br>(0.0%) | 0<br>(0.0%) | 0<br>(0.0%) | 2<br>(2.82%) | 0<br>(0.0%) | 1<br>(1.41%) | 11<br>(15.49%) |
| SSH DMT | 12<br>(16.90%) | 0<br>(0.0%) | 0<br>(0.0%) | 0<br>(0.0%) | 0<br>(0.0%) | 0<br>(0.0%) | 0<br>(0.0%) | 12<br>(16.90%) |
| ATBUTH Bauchi | 7<br>(9.90%) | 0<br>(0.0%) | 0<br>(0.0%) | 1<br>(1.41%) | 1<br>(1.41%) | 0<br>(0.0%) | 0<br>(0.0%) | 9<br>(12.68%) |
| FMC Gombe | 7<br>(9.90%) | 0<br>(0.0%) | 0<br>(0.0%) | 0<br>(0.0%) | 2<br>(2.82%) | 0<br>(0.0%) | 0<br>(0.0%) | 9<br>(12.68%) |
| FMC Jalingo | 9<br>(12.68%) | 2<br>(2.82%) | 1<br>(1.41%) | 1<br>(1.41%) | 0<br>(0.0%) | 1<br>(1.41%) | 0<br>(0.0%) | 14<br>(19.72%) |
| Total (%) | 58<br>(81.6%) | 2<br>(2.8%) | 1<br>(1.4%) | 3<br>(4.23%) | 5<br>(7.04%) | 1<br>(1.41%) | 1<br>(1.41%) | 71<br>(100.0%) |
| Single Gene=61 (85.92%) |  |  |  | Multiple Gene=10 (14.08%) |  |  |  |  |

Key: SSH=State Specialist Hospital; FMC=Federal Medical Center; ATBU-TH=Abubakar Tafawa Balewa Univeristy Teaching Hospital; ESBL=Extended Spectrum Beta-Lactamase.

**Table 6.0.** Summary Table of Molecular Findings Targeted Gene Function Amplicon Size Regional Hotspot Key Observation.

| Targeted Gene | Function | Amplicon Size | Regional Hotspot | Key Observation |
| --- | --- | --- | --- | --- |
| *bla*<sub>TEM</sub> | Penicillinase | 868 bp | Ubiquitous | Found in all six states; most mobile. |
| *bla*<sub>SHV</sub> | Beta-lactamase | 865 bp | Jalingo | High diversity in Taraba State. |
| *bla*<sub>CTX-M</sub> | Cefotaximase | 499 bp | Yola / Gombe | Strongest link to cephalosporin failure |

**Figure 2:**
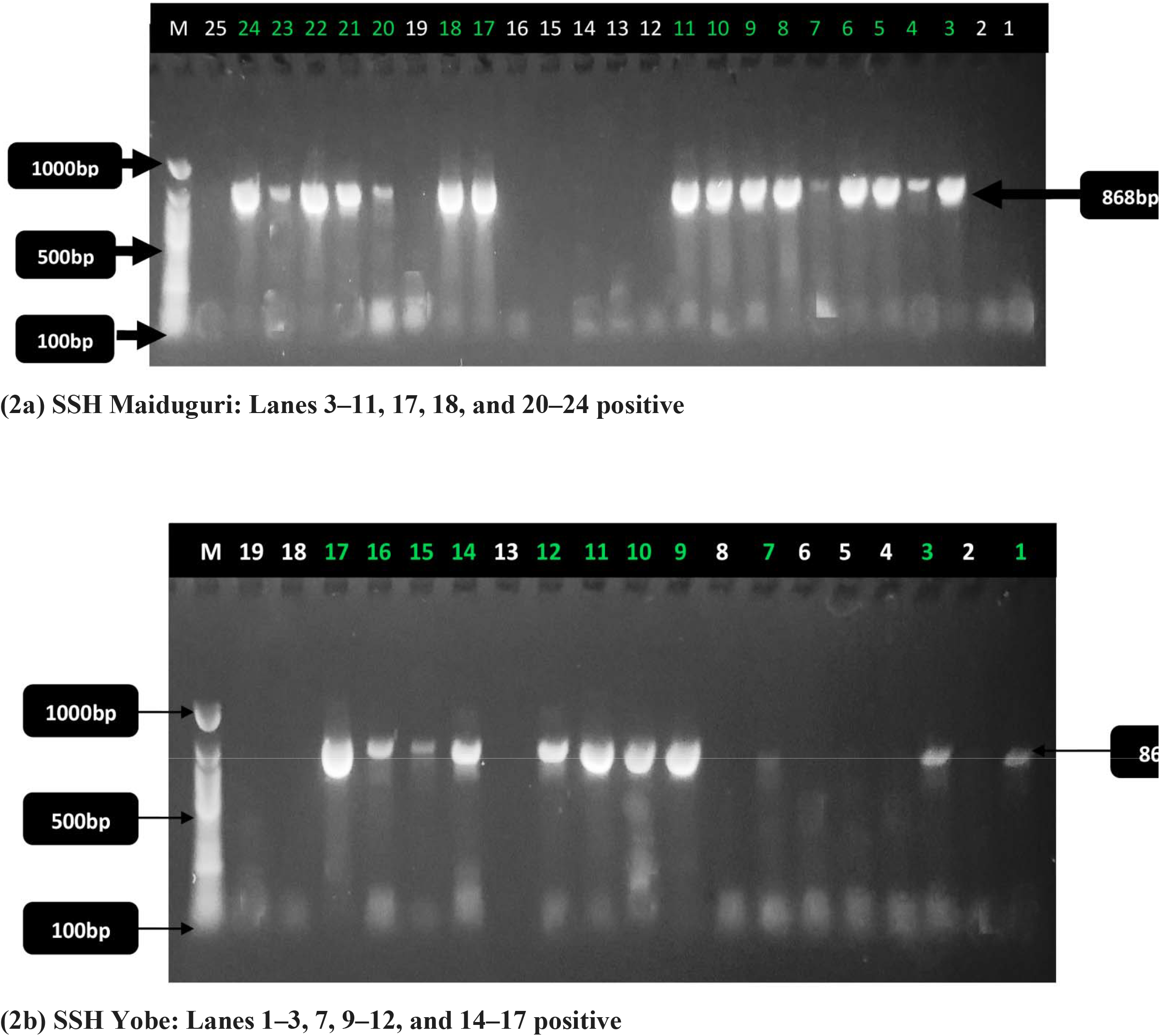
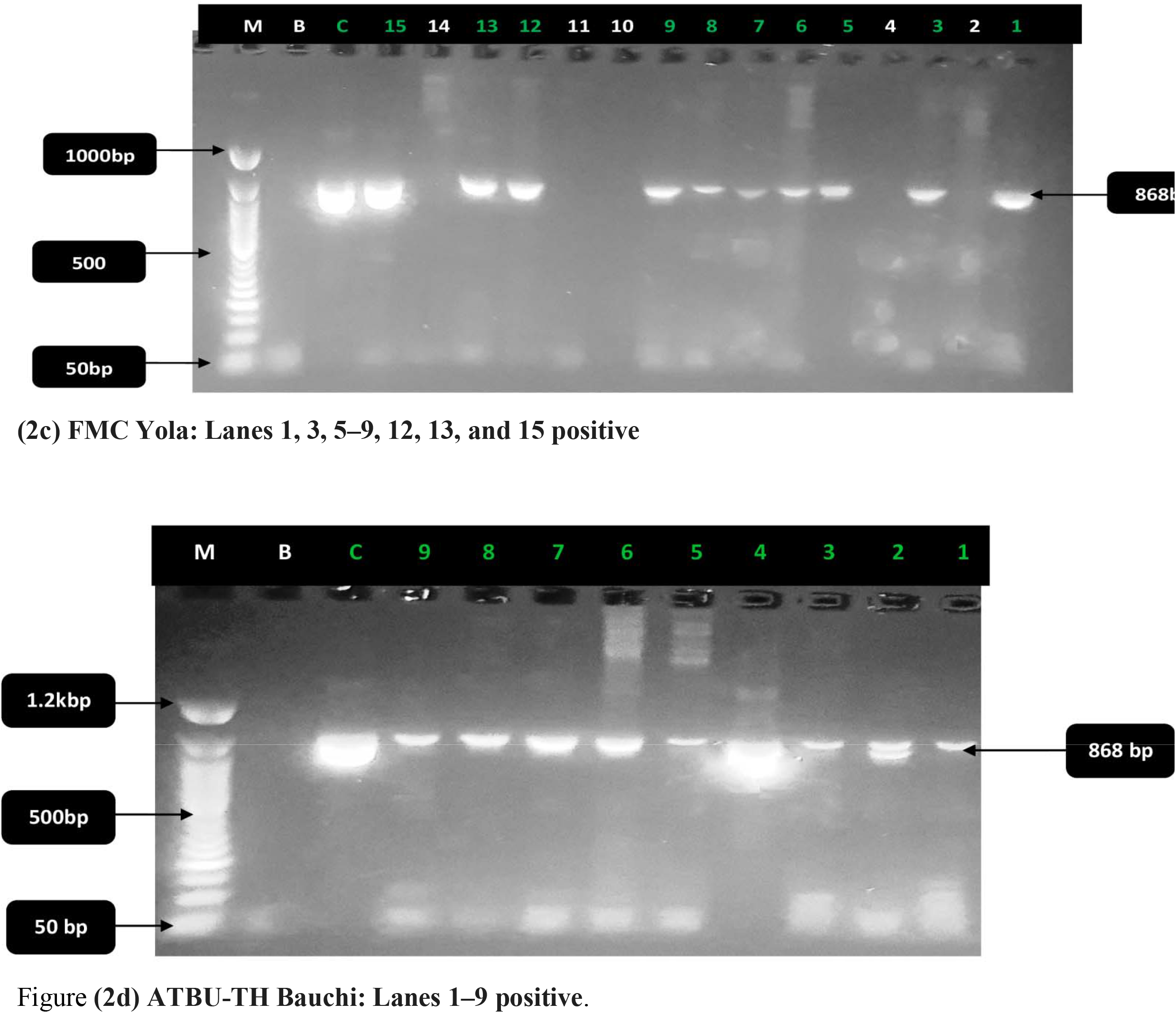
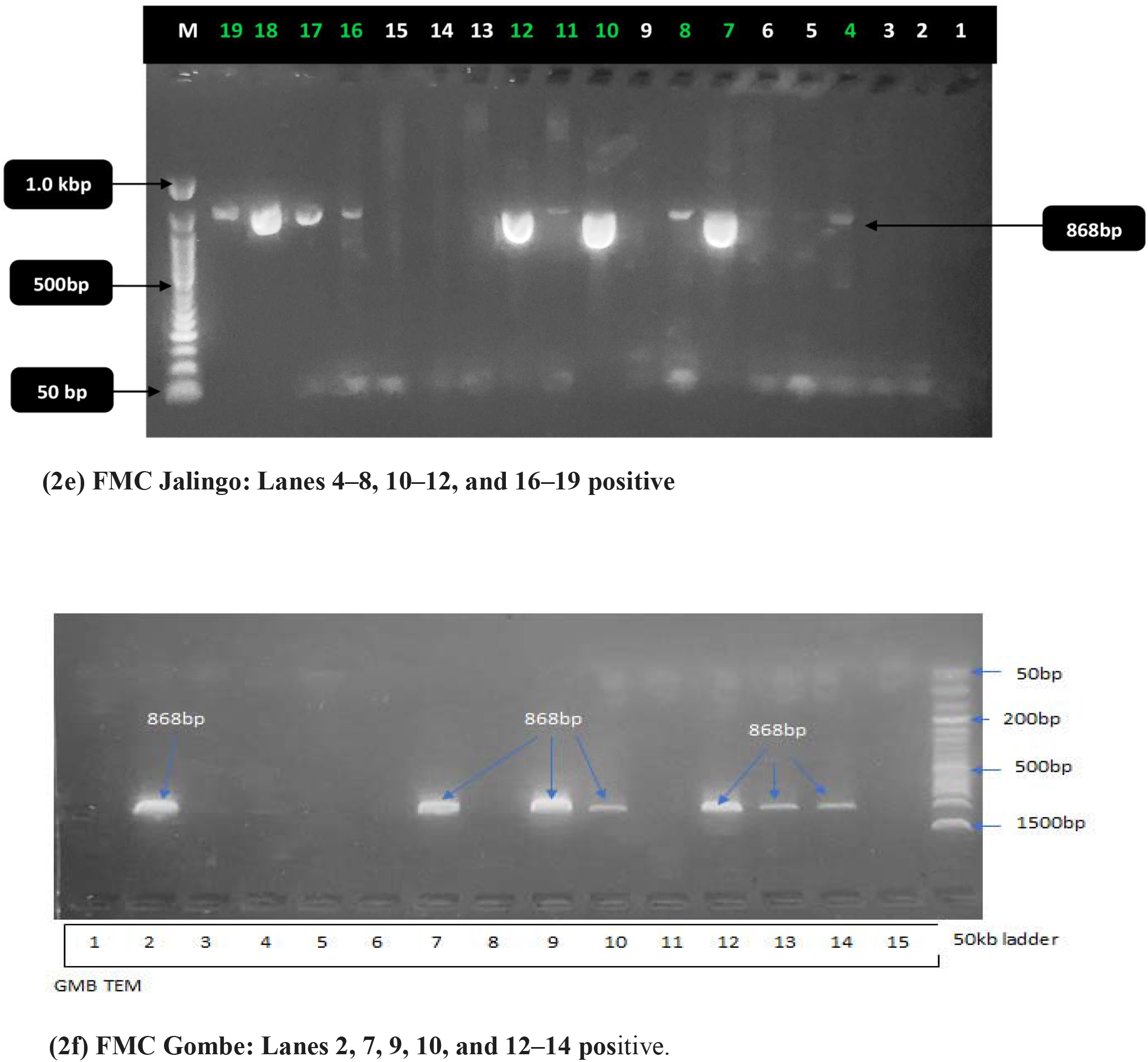
Regional Distribution of the *bla*TEM Genotype (868 bp) in Clinical Proteus Isolates. This figure presents representative images from agarose gel electrophoresis (1.5%) illustrating PCR amplicons for *bla*TEM across six states in North-Eastern Nigeria. M: DNA ladder (50 bp or 100 bp, as indicated); B: blank/Negative control; C: Positive control. The distinct bands observed at 868 bp indicate the presence of positive isolates.

**Figure 3:**
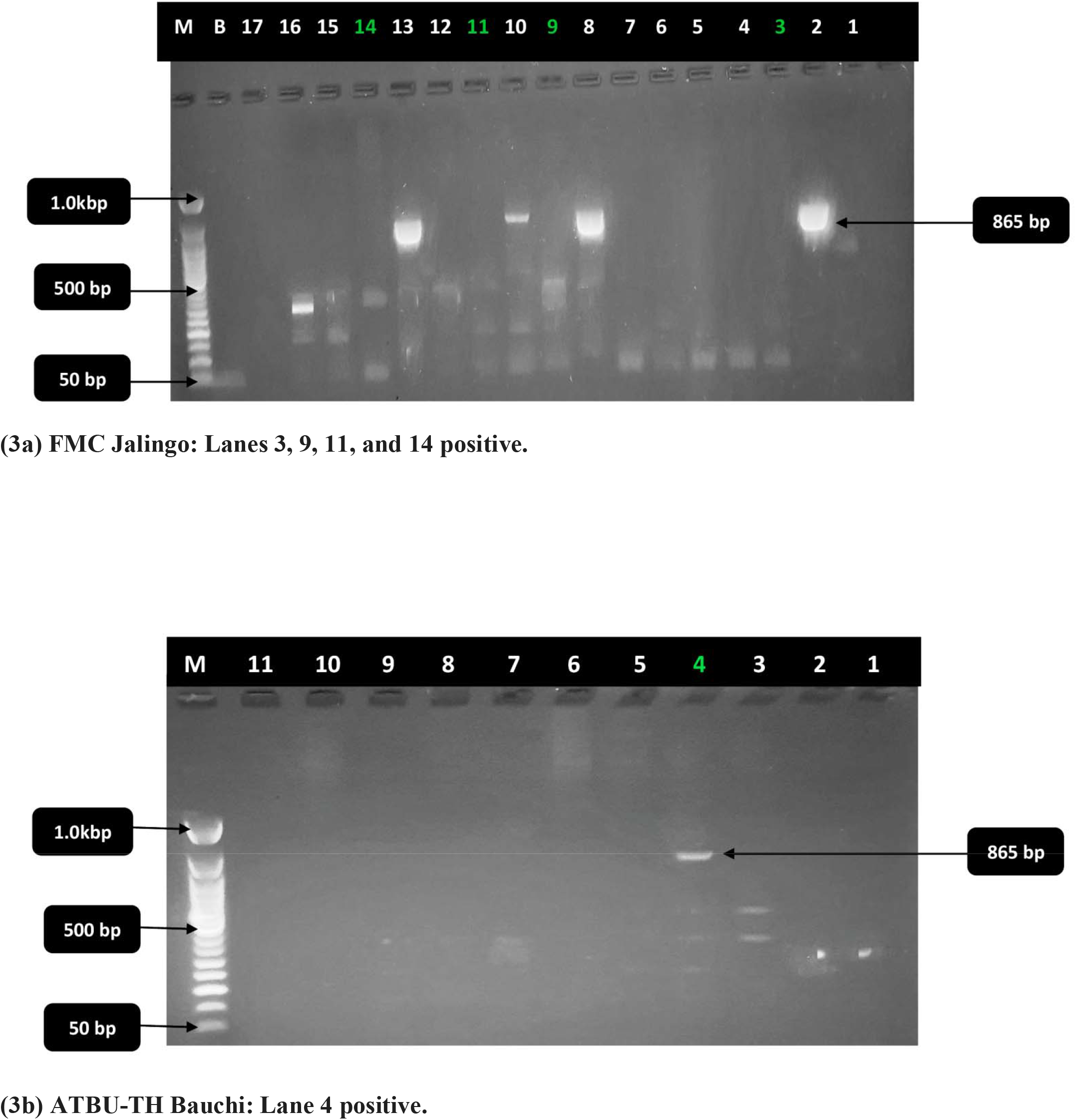
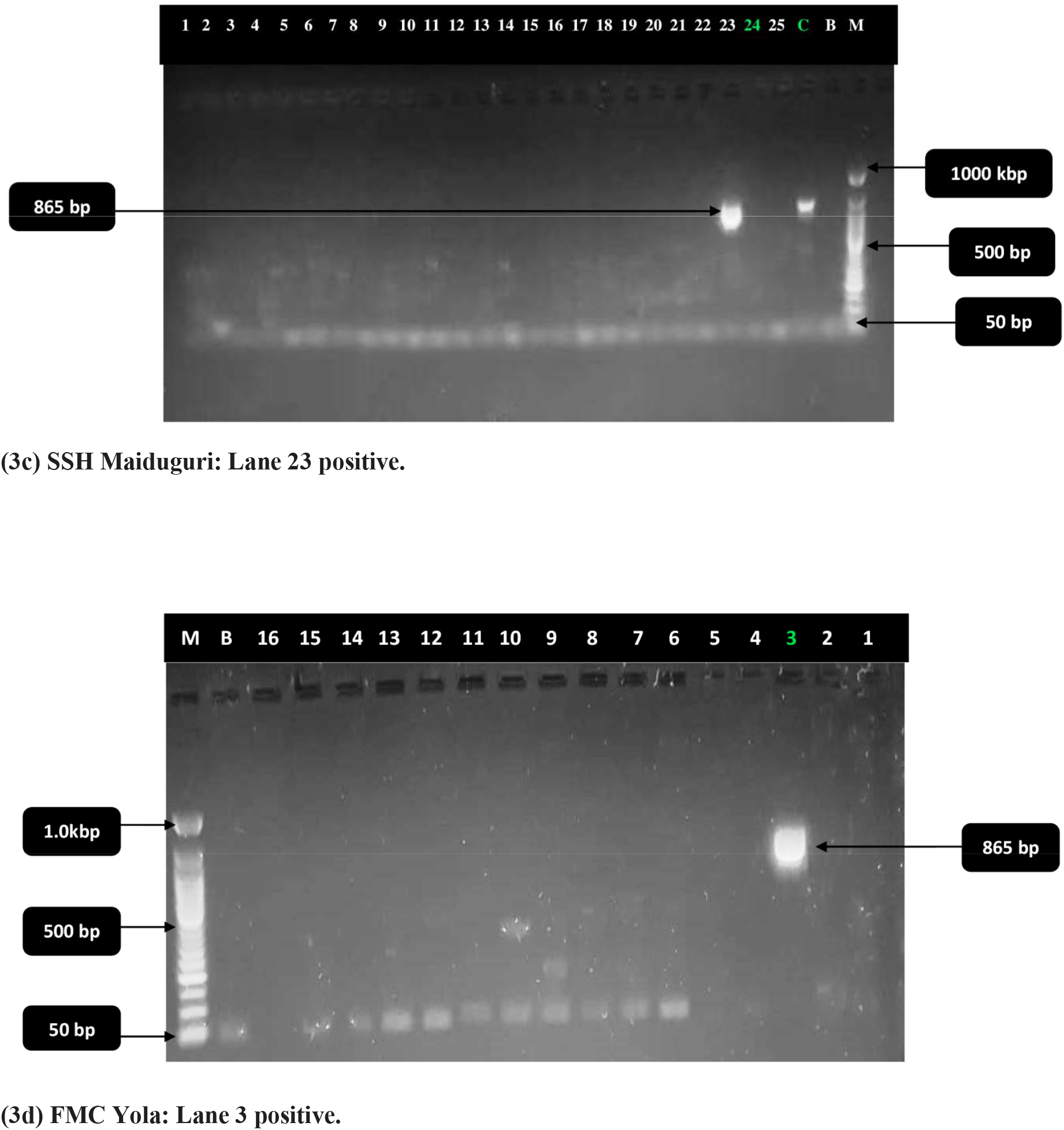
Molecular Identification of the *bla*SHV Genotype (865 bp) in Designated Study Locations. The images obtained from agarose gel electrophoresis (1.5%) serve to confirm the presence of the *bla*SHV gene. M denotes the 50 bp DNA ladder; B represents the negative control; and C indicates the positive control. Positive amplicons are observable at 865 bp

**Figure 4:**
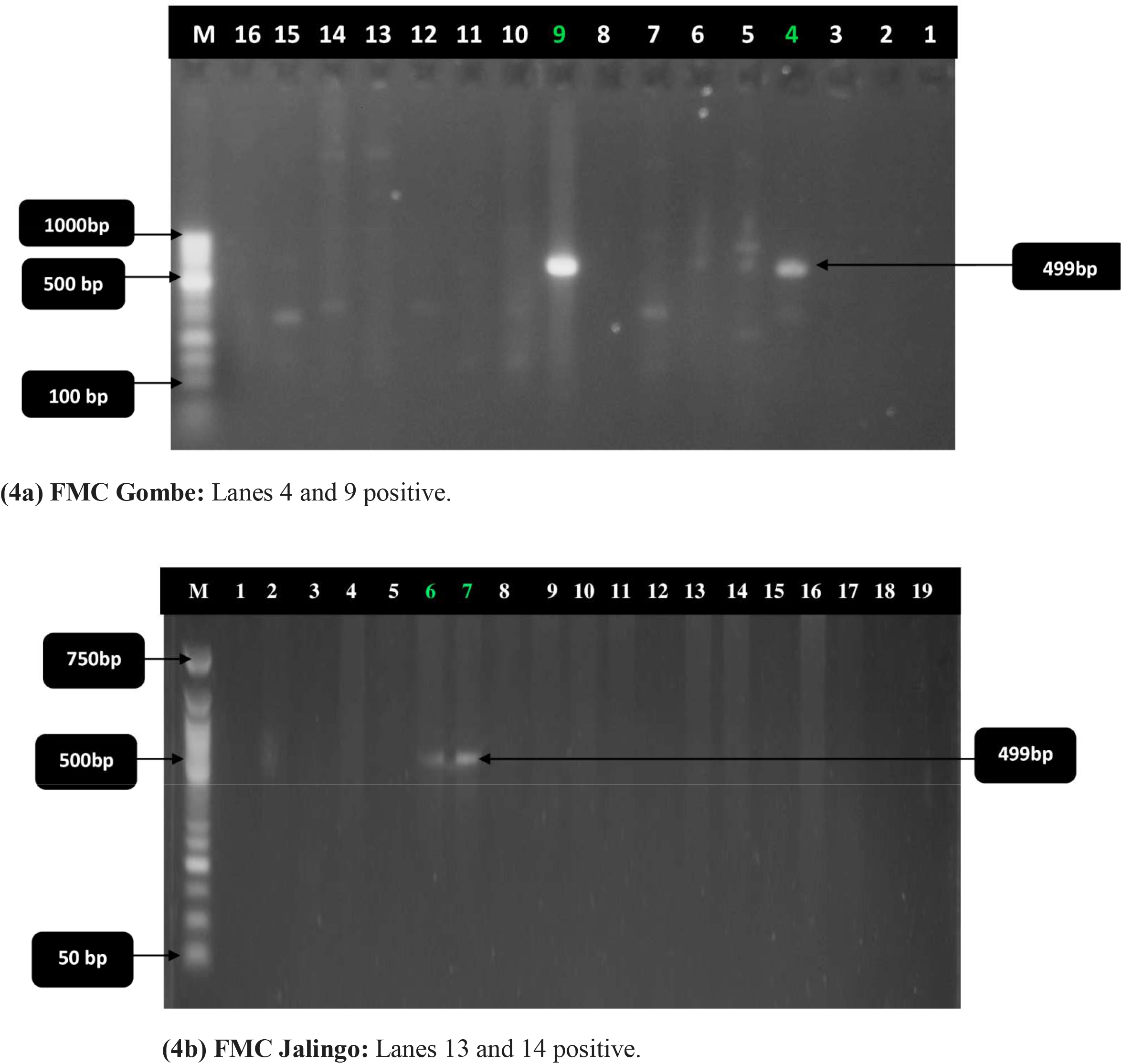
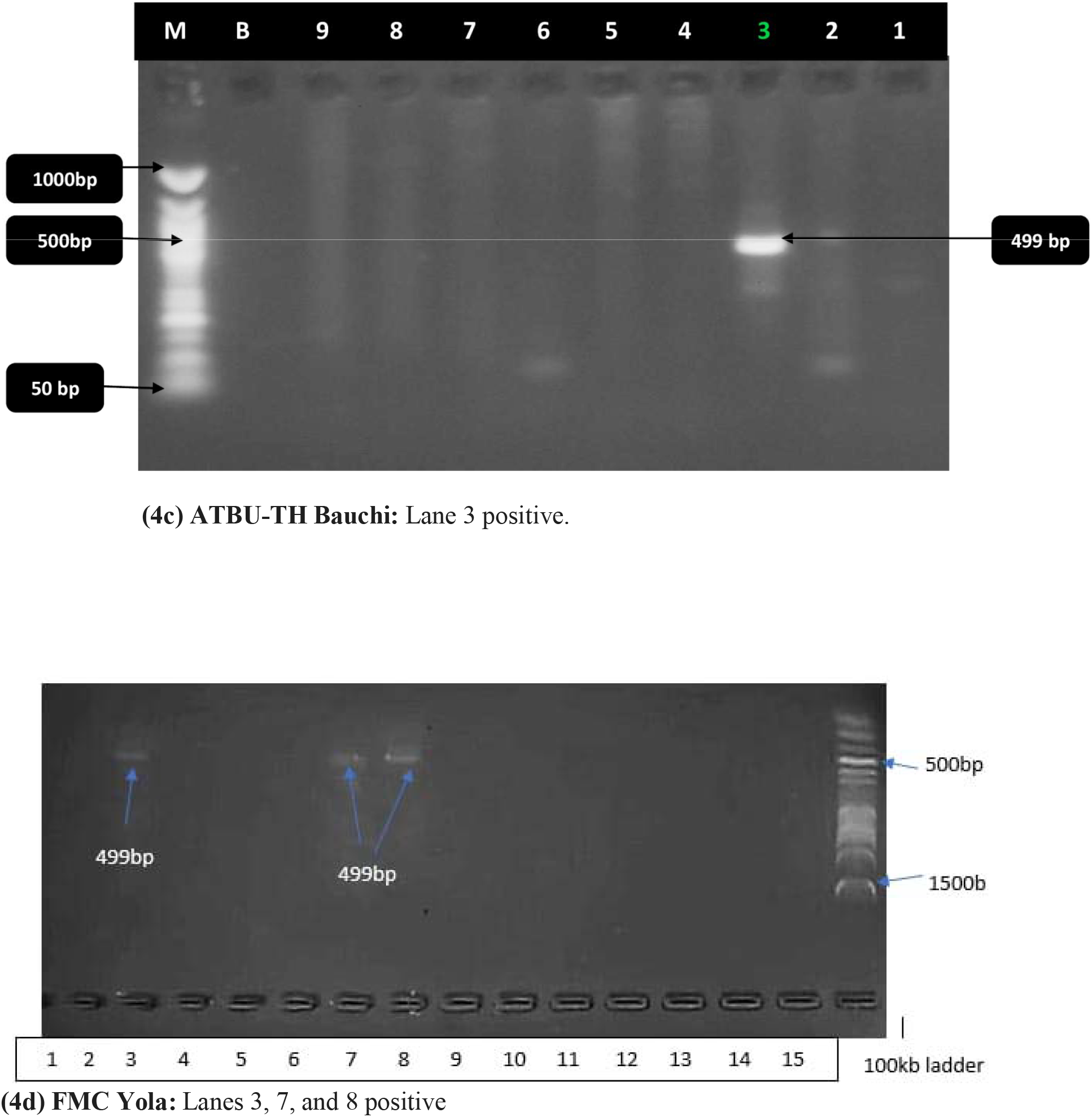
Molecular Identification of the *bla*CTX-M Genotype (499 bp) within the North-Eastern Clinical Corridor. The images illustrated from agarose gel electrophoresis (1.5%) demonstrate the distribution of the *bla*CTX-M gene. M signifies the 50 bp or 100 bp DNA ladder. Arrows highlight the band of interest at 499 bp.

- The *bla*TEM Genotype (868 bp): As the most widely distributed marker, comprising 81.69%, the *bla*TEM genotype was detected across all six states. Distinct amplicons at the 868 bp position were observed in the regional referral centers located in Maiduguri (Fig. 2a), Yobe (Fig. 2b), Yola (Fig. 2c), Bauchi (Fig. 2d), Jalingo (Fig. 2e), and Gombe (Fig. 2f).
- The *bla*SHV Genotype (865 bp): This marker exhibited a targeted presence at 865 bp, particularly in FMC Jalingo (Fig. 3a), which displayed the highest diversity with four positive isolates. Additionally, it was confirmed in Bauchi (Fig. 3b), Maiduguri (Fig. 3c), and Yola (Fig. 3d).
- The *bla*CTX-M Genotype (499 bp): Associated with significant cephalosporin resistance, the *bla*CTX-M genotype was identified at 499 bp in Gombe (Fig. 4a), Jalingo (Fig. 4b), Bauchi (Fig. 4c), and Yola (Fig. 4d).

## 4.0 DISCUSSION

The findings of the six-state regional surveillance study underscore a significant and evolving public health threat in North-Eastern Nigeria. The overall prevalence of *Proteus species*, recorded at 9.60%, aligns with recent multi-center reports from South-Western Nigeria (8.2%) and Ghana (10.1%) [11, 12]. Nonetheless, the predominant presence of *P. mirabilis*, which accounts for 90.97%, indicates a global trend in which this species is outperforming others in clinical settings, attributed to its enhanced biofilm-forming capabilities [4, 13]. The identification of *P. penneri* in Bauchi, with a prevalence of 0.69%, reflects emerging reports from high-acuity units in Europe, suggesting that the region is now hosting high-risk lineages that were previously confined to more developed healthcare systems [14].

A critical contribution of this study is its alignment with the World Health Organization’s Global Antimicrobial Resistance and Use Surveillance System (GLASS) framework, which places particular emphasis on monitoring ESBL-producing *Enterobacteriaceae* as a primary indicator of global antimicrobial resistance (AMR) trends [24]. The observed resistance rates to Cefotaxime (80.56%) and Ceftazidime (73.61%) in *Proteus* species significantly exceed the median resistance levels reported in the most recent GLASS regional summary for Africa [17]. This finding suggests that the clinical corridor in North-Eastern Nigeria qualifies as a “high-prevalence” zone, warranting urgent inclusion in the global surveillance network to prevent the international dissemination of these resistant lineages.

Moreover, the confirmed production rate of extended-spectrum beta-lactamases (ESBL) at 69.44% is substantially higher than the 45.3% reported in Lagos in 2023 and the 38% observed in recent surveillance conducted in Kenya [15, 16]. This disparity implies that the Lake Chad Basin is experiencing unique selective pressures, likely exacerbated by porous borders and unregulated access to antimicrobials. Additionally, the detection of Ertapenem resistance at 54.86% (P = 0.011) constitutes a pivotal finding within the framework of the WHO AWaRe (Access, Watch, Reserve) classification. Given that carbapenems are designated as “Reserve” antibiotics, this elevated resistance level indicates a significant breach in the regional antimicrobial hierarchy, reflecting concerning trends observed in South Asia but infrequently documented within sub-Saharan Proteus populations [18, 25].4.3.

### 4.1 Molecular Drivers and Regional Hotspots

The finding that *bla*TEM (81.69%) serves as the predominant driver contrasts with European data, wherein *bla*CTX-M-15 is nearly universally recognized as the marker [19]. However, these results are consistent with findings from North Africa and India, where *bla*TEM continues to represent a robust foundation for multi-drug resistance [20]. The clustering of *bla*SHV in Jalingo and *bla*CTX-M in Yola/Gombe suggests that North-Eastern Nigeria is a mosaic of localized “evolutionary hotspots,” likely shaped by local prescribing practices and deficiencies in hospital-specific infection control measures.

### 4.2 High-Frequency HGT: Proving the Genetic Reservoir

The conjugation success rate of 76.92% constitutes a significant finding in the context of global dissemination of antimicrobial resistance (AMR). Typical conjugation frequencies for Enterobacteriaceae reported in the literature range from 30% to 55% [21, 22]. Our results nearly double these figures, demonstrating that the R-plasmids present in North-Eastern Nigeria exhibit extraordinary horizontal mobility. By successfully transforming the susceptible recipient E. coli J53, we have empirically established that Proteus species in this region are not merely “end-point” pathogens, but rather active contributors to the microbial ecosystem. This phenomenon of “genetic spillover” directly enhances the resistance profiles of WHO Critical Priority Pathogens, positioning Proteus as a principal driver of the AMR crisis in sub-Saharan Africa [3, 14].

### 4.3 Study Limitations and Future Perspectives

Despite the regional scope of the study, certain limitations must be acknowledged. The reliance on conventional polymerase chain reaction (PCR) methods restricted our capacity to identify specific allelic variants or novel carbapenemase genes. Additionally, the absence of Whole Genome Sequencing (WGS) limits our understanding of the precise evolutionary lineages of these mobile elements. Nevertheless, the high frequency of horizontal gene transfer observed strongly indicates that these genes are circulating within the environment and livestock, thereby necessitating a “One Health” investigative approach [8, 23].

### 4.4 Conclusion of the Discussion

**I**n conclusion, this regional surveillance provides empirical data demonstrating that *Proteu*s species in North-Eastern Nigeria have progressed from opportunistic clinical isolates to high-efficiency “Genetic Hubs” for multidrug resistance. The principal finding of this study—a horizontal gene transfer (HGT) rate of 76.92%—represents a significant regional anomaly, nearly doubling many global reports concerning enteric pathogens [21, 22].

By illustrating the rapid transfer of *bla*TEM and *bla*CTX-M into Escherichia coli J53, this research confirms that the Lake Chad Basin constitutes a vital “evolutionary hotspot,” wherein resistance is not restricted to specific species but is rather disseminating fluidly across various taxonomic boundaries. According to the WHO Global Antimicrobial Resistance and Use Surveillance System (GLASS) framework, the pronounced prevalence of resistance to “Reserve” antibiotics, such as Ertapenem (54.86%), indicates the imminent prospect of a therapeutic dead-end. In the absence of a coordinated, regional molecular surveillance initiative to mitigate this horizontal genetic transmission, the clinical landscape in sub-Saharan Africa may face the irreversible establishment of pan-resistant enteric lineages, thereby endangering the future of \beta-lactam chemotherapy [24, 25].

## Data Availability

The data sets generated and analyzed in the current research can be requested from the corresponding author.

## DECLARATIONS

### Ethical Approval and Consent to Participate

The study protocol was executed in strict accordance with the ethical principles outlined in the Declaration of Helsinki. Multi-center ethical clearance and approval for the research were obtained from the appropriate State Ministry of Health Headquarters and hospital Ethics Research Committees throughout North-Eastern Nigeria. Specifically, formal approvals were granted by:

- Borno State: The Ministry of Health, Maiduguri (MOH/GEN/6679/1) and the State Specialist Hospital Maiduguri (SSH/GEN/641/Vol. 1).
- Yobe State: The Hospitals Management Board, Damaturu (YSSH/DTR/GEN/302).
- Gombe State: The Ministry of Health Headquarters, Gombe (MOH ADM/621/VOL. I/243).
- Bauchi State: The Ministry of Health, Bauchi (Protocol Registration No: BSMOH/REC/31/2021; Approval No: NREC/03/11/19B/2021/34).
- Adamawa State: The Federal Medical Centre, Yola (FMCY/HREC/20/96).

In compliance with established international standards for research involving human subjects, written informed consent was secured from all adult participants prior to the collection of samples. For participants under the age of 18, written informed consent was obtained from their parents or legal guardians. All experimental procedures, including the genetic and molecular characterization of Proteus spp., were conducted in adherence to institutional guidelines and the applicable Code for Health Research.

## Consent for Publication

Not applicable. This manuscript does not include any personal data of individuals in any format (such as individual details, images, or videos).

## Authors’ Contributions

IMT and HBA conceptualized and designed the research, carried out the molecular experiments, and prepared the manuscript. UAD, AAI, and MMI managed sample collection and bacteriological analysis at the six regional facilities. AMU and JBU oversaw antibiotic susceptibility testing and phenotypic ESBL screening. HJB executed the statistical analysis and conducted proofreading of the final draft for submission. All authors reviewed and approved the final manuscript.

## Acknowledgments

The authors express their gratitude to the laboratory personnel and management of ATBU-TH Bauchi, SSH Maiduguri, FMC Yola, SSH Damaturu, FMC Jalingo, UMTH, and FMC Gombe for their technical assistance throughout the collection of 1,500 samples.

